# Midlife Measures of General Cognitive Performance in the National Longitudinal Study of Adolescent to Adult Health (Add Health)

**DOI:** 10.64898/2026.06.18.26355806

**Authors:** Allison E. Aiello, Farizah I. Rob, Alden L. Gross, David A. Bennett, Jennifer J. Manly, Brenda L. Plassman, Jennifer Momkus, Kamaryn T. Tanner, Rebecca C. Stebbins, Robert A. Hummer

**Author notes:** Co-first authors (Allison E. Aiello, Farizah I. Rob).

## Abstract

**Objective:** The Add Health Cognitive Assessment, Physical, and Sensory Function Protocol (Add CAPS) was developed to assess cognitive, physical, and sensory function in early midlife in a nationally representative sample in the United States. Using Add CAPS, we developed two general cognitive performance measures.

**Methods:** The sample included 2,525 participants from Add Health Wave VI who completed an in- home assessment of cognitive performance. Confirmatory factor analysis (CFA) was used to derive two general cognitive performance (GCP) scores: (1) a five-domain score based on originally designed cognitive domains (Add CAPS GCP), and (2) a modified score aligned with the Harmonized Cognitive Assessment Protocol (HCAP) framework (Add CAPS GCP-H). We evaluated model fit using Root Mean Square Error of Approximation (RMSEA), Standardized Root Mean Square Residual (SRMR), and Comparative Fit Index (CFI) and tested factor scores for criterion validity.

**Results:** Both models showed good fit (Add CAPS GCP: RMSEA = 0.025, SRMR = 0.031, CFI = 0.968; Add CAPS GCP-H: RMSEA = 0.027, SRMR = 0.033, CFI = 0.962), indicating that they adequately represent the underlying GCP construct.

**Discussion:** The Add CAPS cognitive battery captures a robust, hierarchical structure of GCP across alternative domain specifications. The derived factor scores provide a valuable method for characterizing a person’s cognitive baseline during midlife. Importantly, the Add CAPS GCP-H enhances comparability with the HCAP network, supporting cross-cohort analyses of cognitive aging.

## INTRODUCTION

Alzheimer’s disease and related dementias (ADRD) pose a major public health challenge in the US, with the prevalence expected to rise substantially as the population ages (Hebert et al., 2013). Most cohort studies that include measures of cognitive performance, a key indicator of ADRD risk, focus on samples 65 years and older, when cognitive decline is common (Freedman & Kasper, 2019; Langa et al., 2020). However, emerging evidence suggests that cognitive performance may begin to change as early as the mid-40s (Salthouse, 2009; Singh-Manoux et al., 2012). Precise cognitive measures in early midlife are therefore crucial for detecting subtle early changes and identifying modifiable ADRD risk factors before the onset of overt symptoms (Ferreira et al., 2017; Ritchie et al., 2015).

There remains a scarcity of nationally representative data on cognitive performance among middle-aged adults in the United States, with one nationally representative cohort study of cognition (Midlife in the United States) to date among individuals of average age 55 (Lachman et al., 2014). To fill this gap, the National Longitudinal Study of Adolescent to Adult Health (Add Health) recently introduced the Add Health Cognitive Assessment, Physical, and Sensory (Add CAPS) protocol (Aiello et al., 2026). Add CAPS was developed to measure key aspects of cognitive, physical, and sensory functioning in a US nationally representative sample of 39-51 year-olds with mean age 44 years, addressing the need for additional nationally representative data to understand cognitive aging earlier in the lifecourse than is typically the case (King & Chatterji, 2026).

To design Add CAPS, we selected eleven cognitive measures that assessed a broad range of cognition, with multiple measures representing five cognitive domains (Aiello et al., 2026).

Most of these cognitive domains overlap with those assessed in large national aging cohort studies, notably those of the Harmonized Cognitive Assessment Protocol (HCAP) (Langa et al., 2020), which was developed to enable comparable assessments of cognitive function across studies and countries (Gross et al., 2026; Nichols et al., 2026) Several HCAP studies have published on the factor structure of their cognitive battery by conducting confirmatory factor analysis (CFA) models (Gross et al., 2020, 2025; Jones et al., 2024; Liu et al., 2025). CFA has also been used in other studies as a standard approach for measuring construct validity of cognitive batteries (Lachman et al., 2014; Obhi et al., 2022; Rawlings et al., 2016). Using similar methods as HCAP, we evaluated the construct and criterion validity of the hypothesized factor structure of general cognitive performance underlying the cognitive domains available from Add CAPS measured in a nationally representative cohort of early middle-aged adults living in the US. We generated both domain-specific factor scores and two factor scores representing general cognitive performance (GCP): (1) a measure incorporating all originally designed Add CAPS cognitive measures (Add CAPS GCP), and (2) a modified measure of general cognitive performance (Add CAPS GCP-H) aligned with HCAP’s definition of cognitive domains. Together, these scores provide a baseline measure of midlife general cognitive performance for cross-sectional analyses in Add Health and for tracking cognitive change in future waves. The Add CAPS GCP-H score further supports comparisons with other large-scale studies of cognitive aging in and out of the USA.

## METHODS

### Dataset

Add Health is a nationally representative longitudinal study that has followed adolescents in grades 7-12 in US schools during the 1994-95 academic year through adulthood. Add Health used a school-based clustering sampling design to include a stratified random sample of 20,745 adolescents (Harris et al., 2019; Hummer et al., 2026). Six waves of data collection have been completed to date. In Wave VI, conducted between 2022 and 2025, Add Health participants were between the ages of 39 and 51 with a mean age of 44 years.

Wave VI utilized a two-sample design: Sample 1 (N = 9,366), which was largely web-based data collection, and Sample 2 (N = 2,613), which was largely conducted in-person (Hummer et al., 2025, 2026). The majority of Add CAPS cognitive measures were administered to Sample 2 by field interviewers in participants’ home (Aiello et al., 2026). We used data from Sample 2 in this study.

### Cognitive Measures

Details on Add CAPS cognitive measures have been published previously, and included:: a subset of measures from the TestMyBrain (TMB, a digital neuropsychology battery) (Aiello et al., 2025d); a subset of measures from the NIH Toolbox Cognitive Function Battery (Aiello et al., 2025c); Interviewer-Administered Word Recall and Backward Digit Span (Aiello et al., 2025b); and the Animal Naming Test (Aiello et al., 2025a). The eleven Add CAPS cognitive measures represented five different domains of cognitive performance: executive function, processing speed, verbal episodic memory, working memory, and language/semantic fluency (Aiello et al., 2026).

Measures of executive function included TestMyBrain (TMB) Gradual Onset Continuous Performance Test (GCPT) and NIH Toolbox Dimensional Card Change Sort Test (DCCS). In TMB GCPT, participants were shown images of cities and mountains and instructed to make a keyboard/screen press when cities were displayed and withhold from responding when mountains appeared. The primary outcome is discriminant ability (dPrime), a measure of response accuracy. NIH Toolbox is a matching game with colors and shapes. Participants first matched shapes with a target image, then matched colors. The primary outcome is based on accuracy and reaction time.

Processing speed was assessed through TMB Digit Symbol Matching and NIH Toolbox Pattern Comparison. In the Digit Symbol Matching Test, a legend key of six symbols each associated with a digit 1 through 3 was presented and remained visible throughout the test. During the assessment, individual symbols were presented in a target area, and the participant was instructed to select the corresponding digit as quickly as possible for 90 seconds. The primary outcome was the median reaction time for correct responses. In our analysis, this was reverse-coded to align with all other cognitive assessment outcomes such that higher values indicated better performance. In NIH Toolbox Pattern Comparison, participants were shown two pictures and instructed to respond “yes” if the two pictures were the same or “no” if they were different. The test was conducted either until 130 trials had been displayed, or 85 seconds had passed. The primary outcome was the number of correct responses.

Verbal episodic memory measures included interviewer-administered immediate and delayed word recall, and TMB Verbal Paired Associates. In the interviewer-administered word recall tasks, participants were read a list of 15 words and asked to say as many as they could recall in 90 seconds (immediate). After a delay of approximately 1-2 minutes, participants were given 60 seconds to recall the words again (delayed). In TMB Verbal Paired Associates, participants were visually presented with 25 pairs of words and, after a delay of 1.5-2.5 minutes, they were shown one word at a time and asked to choose the correct paired word from a list of four options. The primary outcome was the accuracy, the proportion of correct responses out of the total responses.

Working memory was assessed through interviewer-administered Backward Digit Span and TMB Backward Digit Span. In the interviewer-administered test, participants were presented with strings of numbers of increasing length (starting with 2 digits up to 8 digits) and asked to recall each backwards. The primary outcome was the length of the longest string correctly recalled. TMB Backward Digit Span was similar, but sequences were presented visually, and number strings ranged from 2 to 11 digits.

Language/semantic fluency was assessed through the interviewer-administered Animal Naming Test and NIH Toolbox Picture Vocabulary Test. Participants were asked to name as many animals as they could in 60 seconds in the Animal Naming Test. In the NIH Toolbox Picture Vocabulary Test, participants were given a word from an audio recording and instructed to select the corresponding image from a choice of four images. The primary outcome of the Picture Vocabulary Test was a derived score based on the participant’s overall ability.

### Sample Description

The full Add CAPS cognitive battery was administered to Sample 2, which served as the primary sample for descriptive characteristics and CFA. Among Sample 2 participants (N = 2,613), 91.1% completed all cognitive measures, while 8.9% were missing between 1 to 11 measures (**Supplementary Table 1**). Sample 2 included oversamples of Black, Hispanic, and Asian/Pacific Islander Add Health participants (Hummer et al. 2025); thus, all analyses incorporated survey weights to make results generalizable to the target population of US midlife adults who were in grades 7-12 of American schools in the 1994-1995 school year (Liao et al., 2025). Participants were included if they completed at least one cognitive measure (n = 11 excluded). An additional 77 participants were removed due to missing survey design variables, resulting in a final analytic sample of 2,525 participants. Two participants with implausibly long median reaction times on the TMB Digit Symbol Matching test were recoded to missing for that assessment. Weighted descriptive statistics were calculated for the analytic sample, and Pearson’s correlation coefficients between the cognitive measures were calculated to assess convergent and discriminant validity.

### Confirmatory Factor Analysis

We conducted CFA models in several steps, presented in **Supplementary Figure 1**. First, we evaluated the fit of single-factor models for each cognitive domain. Second, we evaluated multidimensional correlated factors models to assess the degree of correlation among domains and evaluate whether any domains might be combined based on theoretical and statistical considerations. Third, we specified a second-order factor model in which general cognitive performance was modeled as an underlying construct. This approach allowed us to evaluate the fit of the hypothesized hierarchical structure and derive a general cognitive performance factor score. Finally, we repeated the same steps using an approach that aligned more closely with HCAP’s specification of executive function in their CFA-based measure of general cognitive performance. More details on each model type are below.

In CFA models, all variables were treated as continuous and used min-max normalization to scale each to a range of 0-1 prior to factor analysis. We used the robust maximum likelihood (MLR) estimator, with full-information maximum likelihood to handle missingness based on the conditionally random missingness assumption. CFA models were estimated using Mplus (version 9, Muthén & Muthén, Los Angeles, CA).

### Add CAPS General Cognitive Performance Measure (Add CAPS GCP)

We first fit separate single-factor CFA models to evaluate the fit of each cognitive domain individually. All hypothesized cognitive domains in Add Health consist of 2-3 indicators (cognitive measures). Single-factor models with 2-3 indicators are saturated with zero degrees of freedom and exhibit perfect fit (with additional constraints for 2-indicator factor models) (Brown, 2015).

Next, we conducted multidimensional CFA models to examine the correlations between the five cognitive domains. To improve model fit, we also specified a model that allowed residual errors of the immediate and delayed word recall tests to be correlated as a means of capturing the shared variance due to methodological similarities.

Finally, we fit a second-order CFA model specifying a general cognitive performance latent variable underlying the five cognitive domains of executive function, processing speed, episodic memory, working memory, and language/semantic fluency. This model addressed our goal to assess whether our hypothesized hierarchical structure of cognition adequately explained the correlations between the five cognitive domains.

### Add CAPS General Cognitive Performance Measure- following HCAP (Add CAPS GCP-H)

In contrast to the domain structure used to form Add CAPS GCP, prior studies including the HCAP (Gross et al., 2020, 2025; Lachman et al., 2014) have specified executive function as a broad domain: executive function, processing speed/attention, and working memory measures.

Adopting a similar approach, we first fit a single-factor model for executive function with six indicators (two measures each of executive function, processing speed, and working memory) to develop a measure of executive function that is comparable to HCAP. Next, we conducted a multidimensional CFA model to examine the correlations between the three cognitive domains (the coarse executive function domain, episodic memory, and language/ semantic fluency). Last, we fit a second-order CFA model specifying a general cognitive performance factor underlying these three cognitive domains. This model addressed our aim to create a general cognitive performance measure aligned with the executive function domain used across HCAP studies.

### Model Fit

Model fit was evaluated using the root mean squared error of approximation (RMSEA), confirmatory fit index (CFI), and standardized root mean square residual (SRMR) (Hu & Bentler, 1999). RMSEA is as an absolute fit index that rewards more parsimonious models; CFI is a comparative fit index that tests the improvement of the model over a null model (Hu & Bentler, 1999); SRMR is an index of absolute fit assessing the average standardized difference between model-implied and observed covariances. Following practices from the HCAP studies, we used the following criteria to assess fit: perfect fit if CFI = 1, and RMSEA = 0, and SRMR = 0; good fit if CFI >= 0.95, and RMSEA <= 0.05, and SRMR <= 0.05; adequate fit if CFI >= 0.90, and RMSEA <= 0.08, and SRMR <= 0.08; poor fit if CFI < 0.90, or RMSEA > 0.08, or SRMR > 0.08. We computed the coefficient omega (ω) and second-order omega coefficient, estimates of internal consistency, from CFA models. For single-factor models, omega was calculated using the sum of squared factor loadings divided by the sum of squared factor loadings and residual variances of indicator variables (Peters, 2018). Computation of second-order omegas additionally included residual variances of first-order factors. We considered omega values >= 0.70 to be satisfactory (Nunnally & Bernstein, 1994).

### Factor Scores and Criterion Validity

Regression method factor scores were estimated for the general cognitive performance and cognitive domains from adequate to well-fitting models using Mplus, along with the standard error of measurement for the factor scores. Since our CFA models consisted of all continuous indicators, the standard error of measurement was not person-specific but rather dependent on the missingness pattern in indicators.

We tested criterion validity by correlating the general cognitive performance and cognitive domain factor scores with variables known to be related to cognitive performance, including age, education attainment, self-reported health variables (e.g. Add Health survey measures of memory, health, physical functioning difficulties, and the CES-D depression scale). In addition, we examined correlations between the Add CAPS GCP score and a TMB summary score (average of the four z-scored TMB assessment outcomes). We carried out this analysis because TMB tests are available for all participants in the full Add Health Wave VI cohort (i.e., Sample 1 and Sample 2), rather than exclusively in Sample 2. This analysis allowed us to evaluate whether a z-scored TMB summary score could provide a reasonable proxy for the Add CAPS GCP score in analyses where the full cognition battery was unavailable. Details on the construction of these variables are presented in **Supplementary Table 3**.

## RESULTS

### Descriptive Characteristics

Weighted characteristics of the analytic sample (N=2,525) are presented in **Table 1**. The sample was evenly distributed by sex (49.7% females), with a mean age of 44.4 years (range 40 – 51). The weighted racial/ethnic distribution was 68.6% White, 15.6% Black, 10.8% Hispanic, and 3.7% Asian/Pacific Islander. Due to small sample sizes, the “some other race or origin” and “American Indian or Alaska Native” groups were combined into one “other” category (1.3%). Educational attainment was 34.7% college degree or more, 40.8% some college education, and 24.6% with a high school diploma/GED, or less. Survey-weighted means, standard deviations, ranges, and missingness for each cognitive assessment are displayed in **Table 1**. Response rates were high for each cognitive measure (94.9%-97.9%), with domain-specific missingness detailed in **Supplementary Table 2**.

**Table 1.**
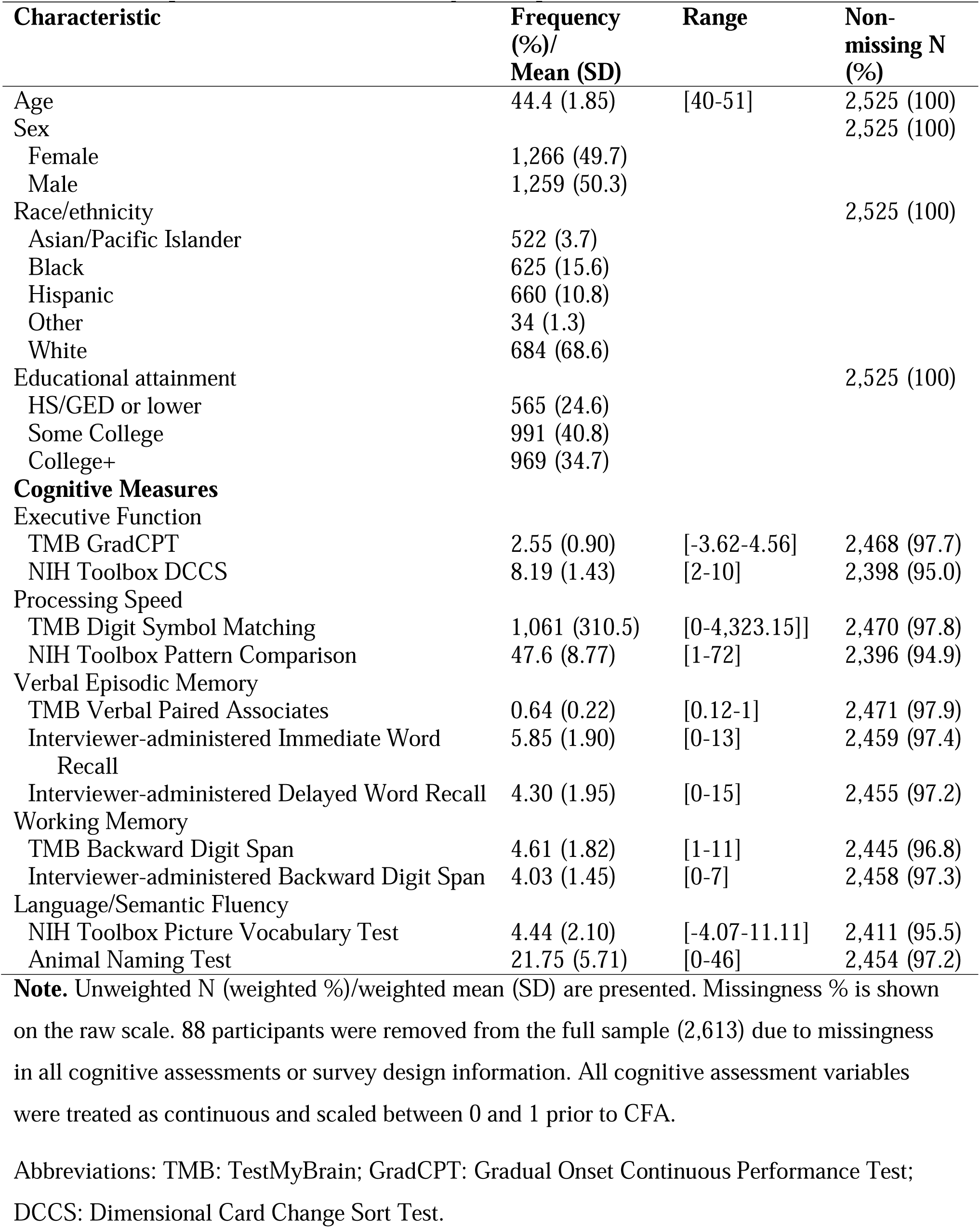
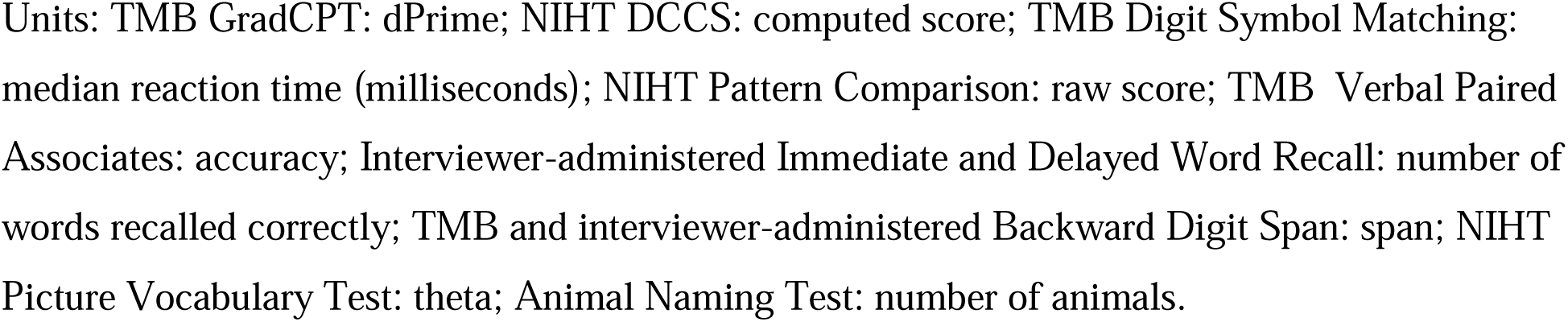
Descriptive characteristics of analytic sample (n = 2,525)

| Characteristic | Frequency<br>(%)/<br>Mean (SD) | Range | Non-<br>missing N<br>(%) |
| --- | --- | --- | --- |
| Age | 44.4 (1.85) | [40-51] | 2,525 (100) |
| Sex |  |  | 2,525 (100) |
| Female | 1,266 (49.7) |  |  |
| Male | 1,259 (50.3) |  |  |
| Race/ethnicity |  |  | 2,525 (100) |
| Asian/Pacific Islander | 522 (3.7) |  |  |
| Black | 625 (15.6) |  |  |
| Hispanic | 660 (10.8) |  |  |
| Other | 34 (1.3) |  |  |
| White | 684 (68.6) |  |  |
| Educational attainment |  |  | 2,525 (100) |
| HS/GED or lower | 565 (24.6) |  |  |
| Some College | 991 (40.8) |  |  |
| College+ | 969 (34.7) |  |  |
| **Cognitive Measures** |  |  |  |
| Executive Function |  |  |  |
| TMB GradCPT | 2.55 (0.90) | [-3.62-4.56] | 2,468 (97.7) |
| NIH Toolbox DCCS | 8.19 (1.43) | [2-10] | 2,398 (95.0) |
| Processing Speed |  |  |  |
| TMB Digit Symbol Matching | 1,061 (310.5) | [0-4,323.15]] | 2,470 (97.8) |
| NIH Toolbox Pattern Comparison | 47.6 (8.77) | [1-72] | 2,396 (94.9) |
| Verbal Episodic Memory |  |  |  |
| TMB Verbal Paired Associates | 0.64 (0.22) | [0.12-1] | 2,471 (97.9) |
| Interviewer-administered Immediate Word Recall | 5.85 (1.90) | [0-13] | 2,459 (97.4) |
| Interviewer-administered Delayed Word Recall | 4.30 (1.95) | [0-15] | 2,455 (97.2) |
| Working Memory |  |  |  |
| TMB Backward Digit Span | 4.61 (1.82) | [1-11] | 2,445 (96.8) |
| Interviewer-administered Backward Digit Span | 4.03 (1.45) | [0-7] | 2,458 (97.3) |
| Language/Semantic Fluency |  |  |  |
| NIH Toolbox Picture Vocabulary Test | 4.44 (2.10) | [-4.07-11.11] | 2,411 (95.5) |
| Animal Naming Test | 21.75 (5.71) | [0-46] | 2,454 (97.2) |

**Note.** Unweighted N (weighted %)/weighted mean (SD) are presented. Missingness % is shown on the raw scale. 88 participants were removed from the full sample (2,613) due to missingness in all cognitive assessments or survey design information. All cognitive assessment variables were treated as continuous and scaled between 0 and 1 prior to CFA.
Abbreviations: TMB: TestMyBrain; GradCPT: Gradual Onset Continuous Performance Test; DCCS: Dimensional Card Change Sort Test.
Units: TMB GradCPT: dPrime; NIHT DCCS: computed score; TMB Digit Symbol Matching: median reaction time (milliseconds); NIHT Pattern Comparison: raw score; TMB Verbal Paired Associates: accuracy; Interviewer-administered Immediate and Delayed Word Recall: number of words recalled correctly; TMB and interviewer-administered Backward Digit Span: span; NIHT Picture Vocabulary Test: theta; Animal Naming Test: number of animals.

**Figure 1** presents survey-weighted Pearson correlations among cognitive measures. Within-domain correlations ranged from 0.26 to 0.72, with the strongest observed for Immediate and Delayed Word Recall (episodic memory; r = 0.72), followed by TMB Backward Digit Span and interviewer-administered Backward Digit Span (working memory; r = 0.55). Moderate correlations were observed for TMB GCPT and NIH Toolbox DCCS (executive function; r = 0.45), TMB Digit Symbol Matching and NIH Toolbox Pattern Comparison (processing speed; r = 0.42), and Animal Naming and NIH Toolbox Picture Vocabulary Test (language/semantic fluency; r = 0.40). TMB Verbal Paired Associates (episodic memory) showed weaker correlations with Immediate and Delayed Word Recall (r = 0.26–0.28).

**Figure 1.**
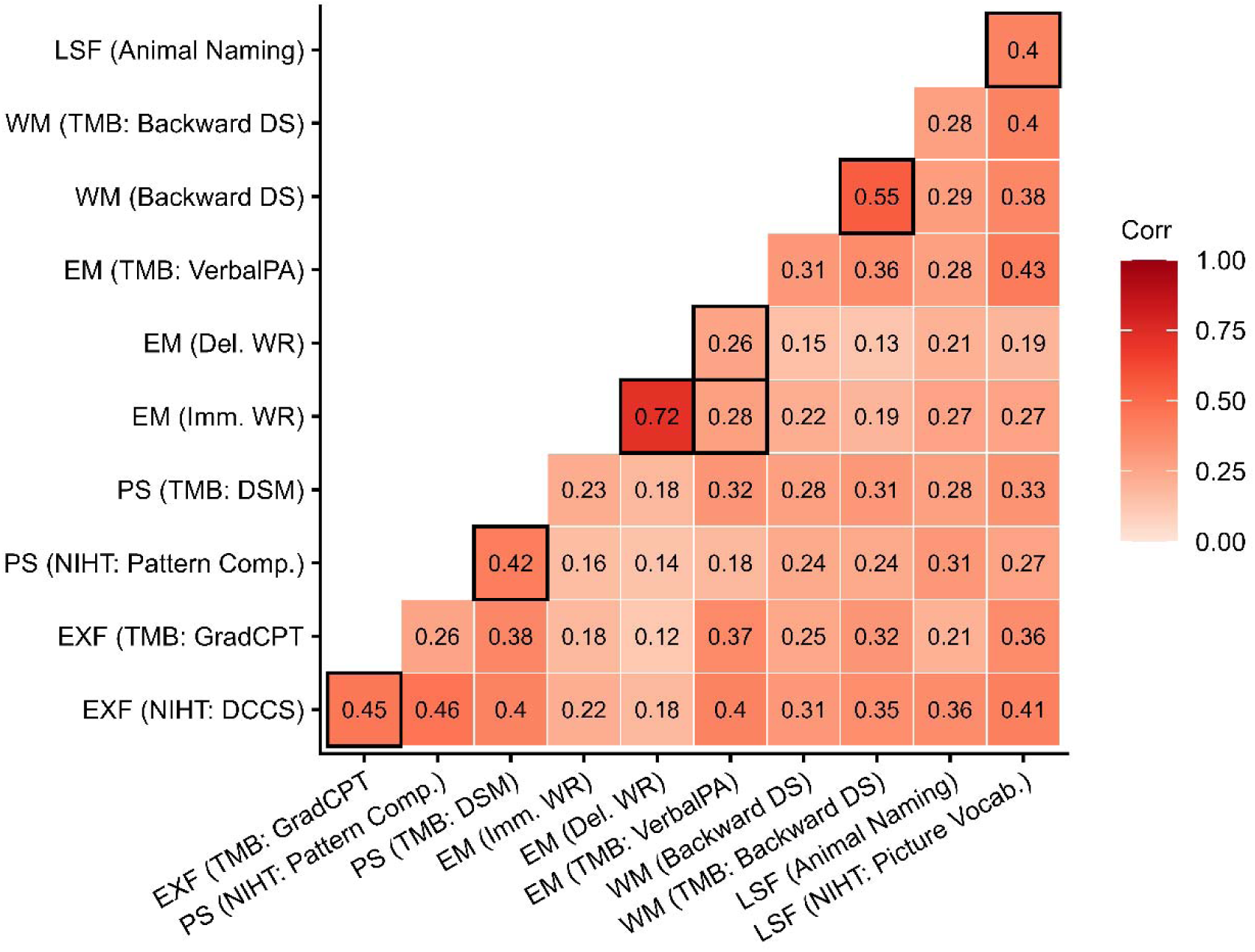
Survey-weighted correlations of individual cognitive assessments (n = 2,525) Pearson’s correlation coefficients are presented. Black boxes are drawn around cognitive assessments hypothesized to measure the same domain. Abbreviations: EXF = Executive Function; PS = Processing Speed; EM = Episodic Memory; WM = Working Memory; LSF = Language/Semantic Fluency; NIHT = NIH Toolbox; TMB = TestMyBrain; DCCS = Dimensional Card Change Sort; GradCPT = Gradual Onset Continuous Performance; DSM = Digit Symbol Matching; WR = Word Recall; DS = Digit Span.

Cross-domain correlations ranged from 0.12 to 0.46 (**Figure 1**). The strongest were between NIH Toolbox DCCS (executive function) and NIH Toolbox Pattern Comparison (processing speed; *r =* 0.46), and between TMB Verbal Paired Associates (episodic memory) and NIH Toolbox Picture Vocabulary Test (language/semantic fluency; *r =* 0.43). Additional moderate to low correlations observed across domains are shown in Figure 1. For example, we observed smaller correlations among measures of episodic memory with processing speed (Immediate Word Recall and NIH Toolbox Pattern Comparison, *r =* 0.16), executive function (Delayed Word Recall and TMB GCPT, *r =* 0.12), and working memory (Delayed Word Recall and Backward Digit Span, *r =* 0.15).

### Confirmatory Factor Analysis

#### Add CAPS GCP

**Table 2** displays fit statistics and reliability estimates of derived CFA models to evaluate the fit of five cognitive domains and Add CAPS GCP. The first set of CFA models examined single-factor models for the five cognitive domains (Models I – V representing executive function, processing speed, verbal episodic memory, working memory, language/semantic fluency, respectively). These single domain models were saturated (degrees of freedom = 0) and showed perfect fit, as expected. The omega coefficient of these five factors ranged between 0.65 and 0.74. Factor diagrams of all single-factor models are shown in **Supplementary Figure 2.**

**Table 2.** Model fit statistics of CFA models: five cognitive domains and Add CAPS GCP.

| Model | Domain | Number of indicators | Omega | SRMR | RMSEA | CFI | Description |
| --- | --- | --- | --- | --- | --- | --- | --- |
| <u>Single-factor models</u> |  |  |  |  |  |  |  |
| I | Executive Function | 2 | 0.68 | 0.000 | 0.000 | 1.000 | * |
| II | Processing Speed | 2 | 0.65 | 0.000 | 0.000 | 1.000 | * |
| III | Verbal Episodic Memory | 3 | 0.74 | 0.000 | 0.000 | 1.000 | * |
| IV | Working Memory | 2 | 0.72 | 0.000 | 0.000 | 1.000 | * |
| V | Language/Semantic Fluency | 2 | 0.67 | 0.000 | 0.000 | 1.000 | * |
| <u>Correlated factors models</u> |  |  |  |  |  |  |  |
| VI | Correlated factors | 11 |  | 0.090 | 0.051 | 0.879 | Poor fit |
| VII | Model VI, allowing correlated residual errors of immediate and delayed WR | 11 |  | 0.025 | 0.021 | 0.980 | Good fit |
| <u>Second-order factor model</u> |  |  |  |  |  |  |  |
| VIII | Second-order factor model: Add CAPS GCP with five domains | 11 | 0.74 | 0.031 | 0.025 | 0.968 | Good fit |

**Note.** \* No identified issues other than 2/3 indicators.
Abbreviations: EXF = Executive Function; PS = Processing Speed; WM = Working Memory; BDS = Backward Digit Span; GCP = General Cognitive Performance

To examine the correlations between the five cognitive domains, we developed a correlated factors model (Model VI); model fit was improved by allowing residual errors of immediate and delayed recall of the same word list to correlate (Model VII). We observed good model fit (RMSEA = 0.021; SRMR = 0.025; CFI = 0.980), and factor loadings ranged between 0.34 and 0.80. Correlations between cognitive domain factors ranged between 0.58 and 0.88. The highest correlations were observed between language/semantic fluency and episodic memory (*r* = 0.88) and between executive function and processing speed (*r* = 0.87). Factor diagrams of these two models can be found in **Supplementary Figure 3.**

To explore the fit of our hypothesized factor structure of Add CAPS GCP, we developed a second-order factor model with GCP and five first-order cognitive domain factors (Add CAPS GCP; Model VIII) (**Figure 2**). This model showed good fit (RMSEA = 0.025; SRMR = 0.031; CFI = 0.968). Standardized factor loadings between cognitive domain factors and indicators (cognitive measures) ranged between 0.34 and 0.81. Loadings between cognitive domains and general cognitive performance ranged between 0.73 and 0.92, with executive function and language/semantic fluency exhibiting the strongest loadings on general cognitive performance (**Figure 2**). The second-order omega coefficient was 0.74.

**Figure 2.**
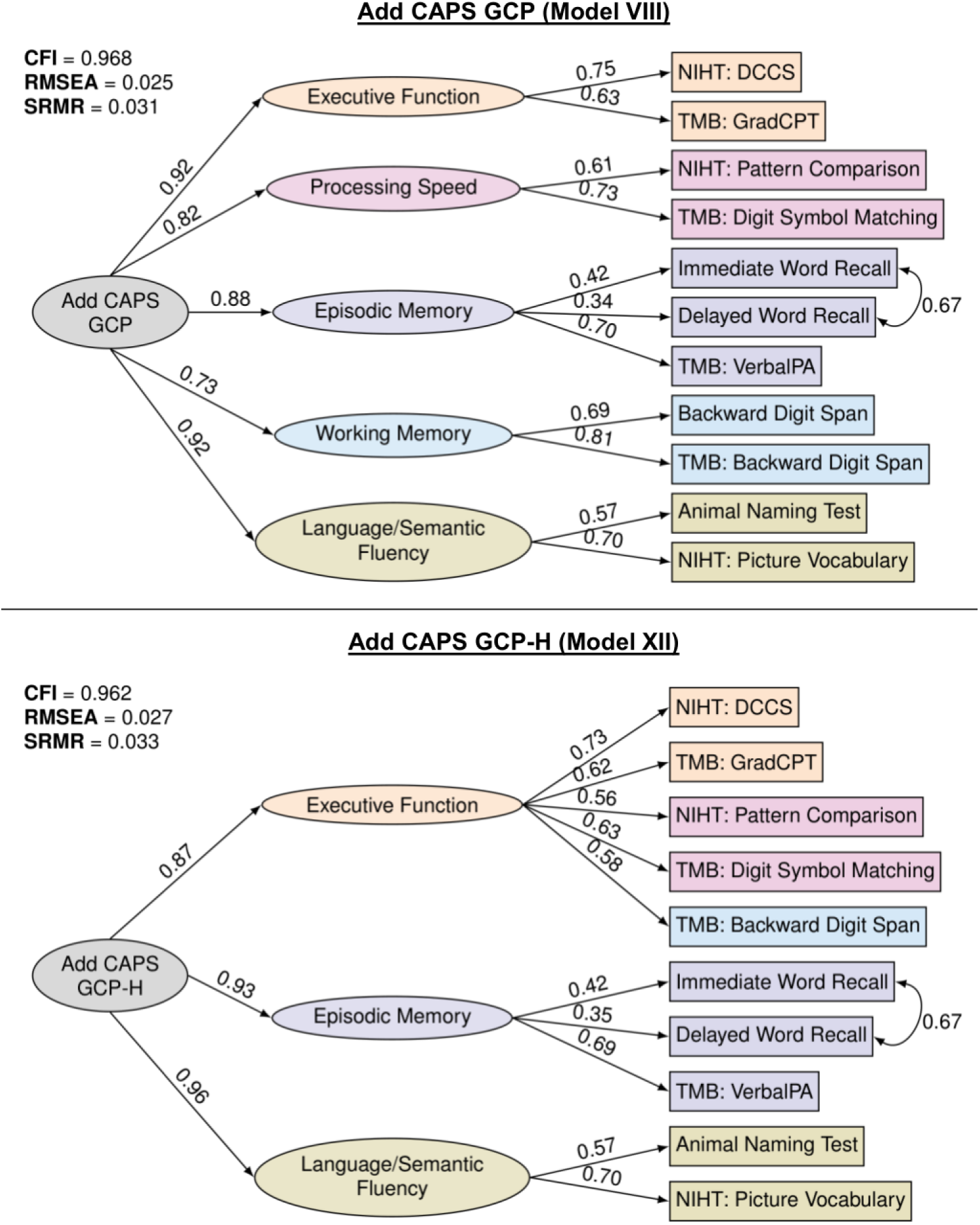
Factor diagrams of second-order CFA models. Standardized factor loadings are presented. Top panel displays the factor diagram of Add CAPS GCP with five cognitive domains (Model VIII). Bottom panel displays the Add CAPS GCP-H with three cognitive domains (Model XII). Abbreviations: NIHT = NIH Toolbox; TMB = TestMyBrain; DCCS = Dimensional Card Change Sort; GradCPT = Gradual Onset Continuous Performance; GCP = General Cognitive Performance.

#### Add CAPS GCP-H

**Table 3** displays fit statistics and reliability estimates of the CFA models, based on the approach used by HCAP studies to specify the executive function domain. To explore the fit of a broad executive function domain comprised of six measures of executive function, processing speed, and working memory, we first conducted a single-factor model (Model IX). This model showed poor fit (RMSEA = 0.065; SRMR = 0.060; CFI = 0.845) and modification indices suggested that the two measures of backward digit span were redundant. Therefore, we fit a model removing interviewer-administered backward digit span (Model X). This model exhibited good fit (RMSEA = 0.032; SRMR = 0.027; CFI = 0.967), and the omega value was 0.51. Factor diagrams of these two models are displayed in **Supplementary Figure 4.**

**Table 3.** Model fit statistics of CFA models: three cognitive domains and Add CAPS GCP-H.

| Model | Domain | Number of assessments | Omega | SRMR | RMSEA | CFI | Description |
| --- | --- | --- | --- | --- | --- | --- | --- |
| <u>Single-factor models</u> |  |  |  |  |  |  |  |
| IX | Executive Function (combining EXF, PS, and WM) | 6 | 0.48 | 0.060 | 0.065 | 0.845 | Poor fit; two BDS tests are redundant |
| X | Executive Function (removing Interviewer-Administered BDS) | 5 | 0.51 | 0.027 | 0.032 | 0.967 | Good fit, but poor reliability |
| <u>Correlated factors models</u> |  |  |  |  |  |  |  |
| XI | Correlated factors (three domains, EXF specified as in Model X) | 10 |  | 0.033 | 0.027 | 0.962 | Good fit |
| <u>Second-order factor model</u> |  |  |  |  |  |  |  |
| XII | Second-order factor model: Add CAPS GCP-H with three domains (EXF modeled as Model X) | 10 | 0.76 | 0.033 | 0.027 | 0.962 | Good fit |

**Note.** Abbreviations: EXF = Executive Function; PS = Processing Speed; WM = Working Memory; BDS = Backward Digit Span; GCP = General Cognitive Performance

To examine the correlations between the three cognitive domains (the broad executive function, episodic memory, and language/semantic fluency), we fit a multidimensional correlated factors model (Model XI). This model also displayed good fit (RMSEA = 0.027; SRMR = 0.033; CFI = 0.962). Episodic memory and language/semantic fluency domains displayed the strongest correlation (*r =* 0.89) (**Supplementary Figure 5**).

To explore whether a factor structure of general cognitive performance underlying three cognitive domains fit our data well, we developed a second-order factor model (Add CAPS GCP-H; Model XII) which displayed good fit (RMSEA = 0.027; SRMR = 0.033; CFI = 0.962). Standardized loadings between cognitive domain factors and indicators (cognitive measures) ranged between 0.35 and 0.73. For the broad domain of executive function, loadings with the five indicators (cognitive measures) were between 0.56 and 0.73. Loadings between executive function, episodic memory, and language/semantic fluency with the general factor were 0.87, 0.93, and 0.96 respectively (**Figure 2**). The second-order omega coefficient was 0.76.

### Factor Scores

In the analytic sample (n = 2,525), 9 participants did not complete any of the five cognitive measures included in the broad executive function domain, and 65 participants did not complete either of the language/semantic fluency measures and were excluded for factor score estimation for these cognitive domains (**Supplementary Table 2**). After assessing model fit and reliability statistics, 5 of the 12 models were selected for estimation of factor scores: episodic memory (Model III, n = 2,525); language/semantic fluency (Model V, n = 2,460); executive function (coarse domain, Model X, n = 2,516); Add CAPS GCP (Model VIII, n = 2,525); and Add CAPS GCP-H (Model XII, n = 2,525).

These factor scores, by design, had an approximate mean of 0. The standard error of measurement associated with the scores was also extracted. The factor scores were standardized to a mean = 0 and standard deviation = 1 for interpretability in standard deviation units, using survey-weighted means and standard deviations. The standard error of measurement was also scaled, to align with the standardized score. The distribution of these factor scores is presented in **Supplementary Figure 6.**

### Criterion Validity and Comparisons with TMB Summary Scores

To evaluate criterion validity, we correlated the general cognitive performance scores (Add CAPS GCP and Add CAPS GCP-H), domain-specific cognitive scores, the TMB Summary Score, self-reported health variables, and age (**Supplementary Figure 7**). For the constructed self-reported scales (**Supplementary Table 3**), higher values indicate poorer self-reported health, memory, and hearing, as well as more physical function difficulties.

Add CAPS GCP and Add CAPS GCP-H scores were nearly perfectly correlated (*r =* 0.99) and showed nearly identical correlations with age (*r* = −0.10), education (GCP: *r =* 0.50; GCP-H: *r =* 0.51) and self-reported health measures, including CES-D (GCP: *r =* −0.14; GCP-H: *r =* −0.13), self-reported health (*r =* −0.06), memory (*r =* −0.15), hearing (*r =* −0.08) and physical function difficulties (*r =* −0.29). Correlations for individual cognitive domains were comparable to those observed for both GCP measures. Add CAPS GCP was strongly positively correlated with the TMB Summary Score (*r =* 0.91), with a slightly lower correlation observed for GCP-H (*r* = 0.87). Among cognitive domains, Executive Function demonstrated the strongest correlation with the TMB Summary Score (*r =* 0.85). Correlations for TMB Summary Score with demographic and health variables were comparable to those observed for GCP measures.

## DISCUSSION

We developed general cognitive performance scores with robust construct and criterion validity for use in Wave VI of Add Health. We derived two confirmatory factor analysis-based scores, Add CAPS GCP and Add CAPS GCP-H, which establish a valuable, precisely characterized midlife cognitive baseline for examining subsequent cognitive decline and identifying early predictors of performance. Importantly, Add CAPS GCP-H provides comparability with the HCAP network, expanding opportunities for complementary cross-cohort analyses of cognitive performance.

Cognitive measures within each domain generally demonstrated expected relationships with one another, providing evidence supporting the construct validity of the underlying cognitive domains. Patterns of correlations among the cognitive measures were broadly consistent with expectations for convergent and discriminant validity. For example, the Picture Vocabulary Test and Animal Naming tests, the two measures of language/semantic fluency, were moderately correlated, while the interviewer-administered immediate and delayed word recall tasks were strongly correlated with one another. These findings suggest that cognitive measures designed to represent the same domains captured shared underlying constructs. However, not all relationships followed the expected patterns. For example, immediate and delayed word recall scores showed lower correlations with the third episodic memory measure, the TMB Verbal Paired Associates assessment. Instead, this assessment showed somewhat stronger correlations with measures from other domains, particularly the NIH Toolbox Picture Vocabulary Test (language/semantic fluency). One possible explanation is that Verbal Paired Associates relies more heavily on associative encoding and cued retrieval processes than free recall tasks, which primarily capture episodic retrieval processes (Naveh-Benjamin, 2000). In addition, paired-associate learning may draw more heavily on semantic knowledge and verbal abilities, which could contribute to stronger associations with vocabulary-based measures. These differences in cognitive demands may result in weaker correlations across these measures despite their classification within the same domain. Differences in task format and mode of administration (interviewer-administered vs. digital self-administered) may also contribute to this pattern (Domingue et al., 2023; Smith et al., 2023). These findings highlight the importance of recognizing potential heterogeneity within broadly defined cognitive domains and suggest that measures categorized under the same domain may capture distinct cognitive processes.

Reliability estimates for individual domains from single-factor models were generally comparable to those observed in other epidemiologic cognitive batteries (Jones et al., 2024; Gross et al., 2025). Episodic memory and working memory showed acceptable internal consistency, whereas executive function, processing speed, and language/semantic fluency exhibited somewhat lower reliability. These differences may reflect the limited number of indicators within some domains or the conceptual heterogeneity of cognitive constructs.

Executive function, for example, encompasses multiple partially overlapping albeit heterogeneous processes important in everyday life, including inhibitory control, cognitive flexibility, and goal-directed behavior, that are difficult to capture with a few brief measures. While the domain-specific scores derived here provide useful summary measures, these findings suggest that they should be interpreted with appropriate caution given the limited number of indicators per domain.

Building on the originally designed Add CAPS domain structure, we evaluated a hierarchical model specifying a general cognitive performance factor underlying five domains. The cognitive domains loaded strongly onto the Add CAPS GCP, indicating substantial shared variance across domains and supporting the interpretation of a general underlying cognitive construct. These findings support the use of a second-order model of general cognitive performance to summarize overall cognitive performance in this cohort. Further, the Add CAPS GCP may be especially useful given this midlife stage, when neurodegenerative processes are generally less advanced than in later life (Hou et al., 2019; Livingston et al., 2020).

In addition to evaluating the originally hypothesized structure, we conducted analyses designed to more closely align with the HCAP-informed specification of executive function. In these analyses, executive function, processing speed, and working memory measures were combined to form a broader executive function domain (Gross et al., 2025, 2020; Jones et al., 2024). We evaluated a three-domain hierarchical model (Add CAPS GCP-H) and the resulting general cognitive performance scores derived from the two hierarchical models were nearly perfectly correlated, indicating that the overall summary measure of cognitive performance was robust to modest differences in domain specification.

Beyond evaluating the structural characteristics of the cognitive battery, we also examined criterion validity by assessing associations between the derived factor scores and survey-based demographic, health and functioning measures. General cognitive performance and cognitive domain factor scores showed associations in the expected directions with age, educational attainment, depressive symptoms, reports of health status, memory difficulties, hearing problems, and physical function limitations. Although correlations with age were modest, this is likely to reflect the relatively narrow and younger age range of participants in the analytic sample rather than modest criterion validity. Overall, the pattern of associations were consistent with established literature linking cognitive functioning to educational attainment, physical and mental health (Clouston et al., 2013; Livingston et al., 2020), providing additional evidence that the latent cognitive scores capture meaningful variation in cognitive performance. Furthermore, the Add CAPS GCP measures were strongly correlated with the average of z-scored TMB tests, indicating that the TMB summary score may provide a reasonable proxy of general cognitive performance and enable analyses in in the full Add Health Wave VI sample, which encompassed both in-person and web-based data collection.

Several limitations should be considered when interpreting these findings. First, the number of cognitive measures representing each domain in the Add CAPS battery was limited, with most domains represented by only two indicators. Although this is common in brief cognitive batteries, the inability to more narrowly estimate domain-specific constructs likely contributed to modest reliability estimates for some domains. The limited number of tests per domain may also adversely influence future measurements of cognitive decline. Second, the confirmatory factor analytic approach used in this study was guided by theory and prior cognitive aging studies, including those using HCAP frameworks; however, alternative factor structures could potentially emerge using exploratory approaches (Liu et al., 2025). For example, instead of focusing on alignment with older populations such as those using HCAP, factor structures that reflect differences in cognitive performance in younger ages may be warranted given the average age of 44 years in Wave VI of Add Health (Agelink Van Rentergem et al., 2020). Third, the Add CAPS battery includes both digitally administered and interviewer-administered cognitive measures. Although our results suggest that these measures can be combined to derive coherent latent constructs, differences in administration mode may still introduce measurement variability related to testing environment, participant familiarity with digital platforms, or other contextual artifacts. Future research could further examine potential mode effects and their implications for cognitive measurement in large population-based studies. Fourth, although Add CAPS GCP-H aligns with HCAP approaches, not all domains included in the original HCAP were available. Future studies should evaluate how well the Add CAPS GCP-H performs in harmonizing existing U.S. and international HCAP cohort data through direct comparison or integration.

The Add CAPS GCP measures establish an important cognitive baseline in Wave VI of Add Health as participants enter midlife, a stage increasingly recognized as critical for understanding the long-term changes underlying cognitive decline and dementia risk. Together, these measures of general cognitive performance and cognitive domains provide a valuable resource for Add Health investigators and create opportunities for work on cognitive function disparities, longitudinal analyses, future opportunities for harmonized comparisons, and integrative studies of cognitive aging and Alzheimer’s disease and related dementias.

## Funding

Data collection and dissemination for Wave VI of Add Health was funded by cooperative agreements U01 AG071448 (PI: Robert A. Hummer) and U01 AG071450 (MPIs: Robert A. Hummer and Allison E. Aiello) from the National Institute on Aging to the University of North Carolina at Chapel Hill, with cooperative funding from five other institutes and offices at the National Institutes of Health: the Eunice Kennedy Shriver National Institute of Child Health and Human Development, the National Institute on Minority Health and Health Disparities, the National Institute on Drug Abuse, the Office of Disease Prevention, and the Office of Behavioral and Social Science Research. Waves I-V of Add Health were funded by grant P01 HD31921 from the Eunice Kennedy Shriver National Institute of Child Health and Human Development, with cooperative funding from 23 other federal agencies and foundations. Add Health is currently directed by Robert A. Hummer at the University of North Carolina at Chapel Hill. Add Health was designed by J. Richard Udry, Peter S. Bearman, and Kathleen Mullan Harris at the University of North Carolina at Chapel Hill. No direct funds from U01 AG071448, U01 AG0-71450, or P01 HD31921 were used in producing this research. Any viewpoints or perspectives expressed in this research are the author’s and do not necessarily represent those of the institutes or agencies that funded Add Health data collection or those of the University of North Carolina at Chapel Hill.

## Conflict of Interest

None declared.

## Data, Codebook, and User Guide Availability

All Wave VI data (e.g., survey, Add CAPS, biological, contextual, birth records, mortality files) have been disseminated in separate files, linkable through an ID number to one another and to previous waves of Add Health. To protect participants, Add Health has developed a tiered dissemination plan. Restricted-use files include all individual participants, physical and biological measures, and administrative and contextual data; they are available to researchers who enter into a contractual agreement with the University of North Carolina – Chapel Hill (UNC-CH). Add Health public-use data files, for which the bar for access is significantly lower, include all survey variables and a limited set of biological variables. To protect against deductive disclosure, the sample size of Add Health public use files is one-third the size of restricted data, with the sample carefully selected back at Wave I. Wave VI public-use data files are available through the Inter-University Consortium for Political and Social Research and National Archive of Computerized Data on Aging (ICPSR/NACDA) at the University of Michigan and through Dataverse at the Odum Institute at UNC-Chapel Hill.

Detailed Wave VI codebooks and user guides are available on the Add Health website. Two types of Wave VI weights are available that account for unequal probability of selection and differential nonresponse: cross-sectional weights address representativeness for estimates that describe the Add Health population within a single wave, while longitudinal weights address analyses that span multiple waves.

Code for this study is not made publicly available; please reach out to corresponding author if you are interested in obtaining the code.

## Data Availability

All Wave VI data (e.g., survey, Add CAPS, biological, contextual, birth records, mortality files) have been disseminated in separate files, linkable through an ID number to one another and to previous waves of Add Health. To protect participants, Add Health has developed a tiered dissemination plan. Restricted-use files include all individual participants, physical and biological measures, and administrative and contextual data; they are available to researchers who enter into a contractual agreement with the University of North Carolina at Chapel Hill (UNC-CH).

## Acknowledgement

Data collection and dissemination for Wave VI of Add Health was funded by cooperative agreements U01 AG071448 (PI: Robert A. Hummer) and U01 AG071450 (MPIs: Robert A. Hummer and Allison E. Aiello) from the National Institute on Aging to the University of North Carolina at Chapel Hill, with cooperative funding from five other institutes and offices at the National Institutes of Health: the Eunice Kennedy Shriver National Institute of Child Health and Human Development, the National Institute on Minority Health and Health Disparities, the National Institute on Drug Abuse, the Office of Disease Prevention, and the Office of Behavioral and Social Science Research. Waves I-V of Add Health were funded by grant P01 HD31921 from the Eunice Kennedy Shriver National Institute of Child Health and Human Development, with cooperative funding from 23 other federal agencies and foundations. Add Health is currently directed by Robert A. Hummer at the University of North Carolina at Chapel Hill. Add Health was designed by J. Richard Udry, Peter S. Bearman, and Kathleen Mullan Harris at the University of North Carolina at Chapel Hill. No direct funds from U01 AG071448, U01 AG0-71450, or P01 HD31921 were used in producing this research. Any viewpoints or perspectives expressed in this research are the author’s and do not necessarily represent those of the institutes or agencies that funded Add Health data collection or those of the University of North Carolina at Chapel Hill. The current study was not preregistered.

## Notes

### Competing Interest Statement

The authors have declared no competing interest.

### Author Declarations

IRB of University of North Carolina at Chapel Hill and Columbia University gave ethical approval for this work.

