## Supplementary Material for "Midlife Measures of General Cognitive Performance in the National Longitudinal Study of Adolescent to Adult Health (Add Health)"

**Section 1: Goals of the study**

**Supplementary Figure 1. CFA Modeling Steps**

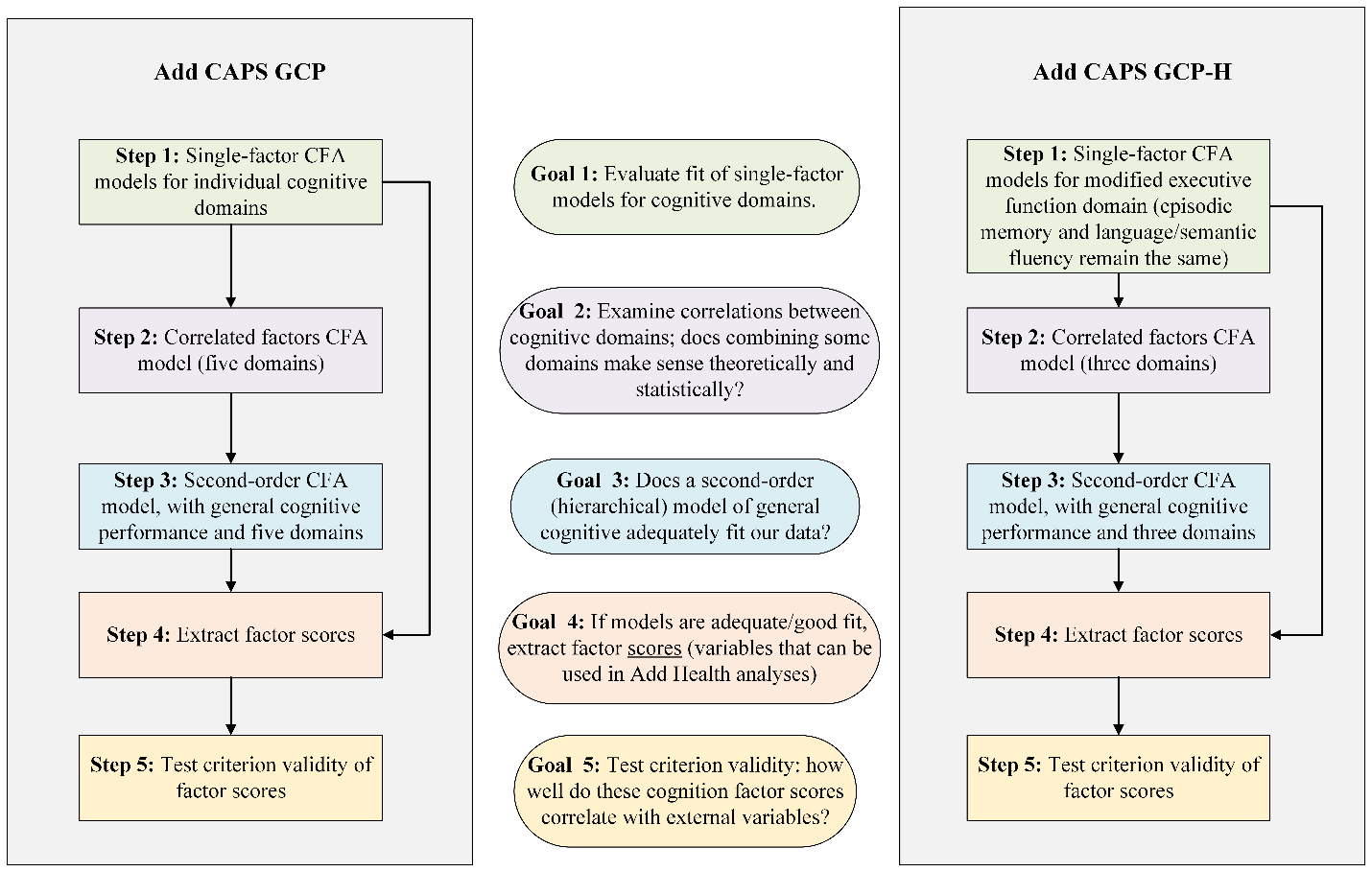

**Section 2: Missingness in Cognitive Assessments**

**Supplementary Table 1. Frequency (%) of participants with missing cognitive assessments**

| **Number of cognitive assessments missing** | **Frequency (N = 2,613)** | **%** |
| --- | --- | --- |
| 0 | 2,380 | 91.1 |
| 1 | 43 | 1.6 |
| 2 | 16 | 0.6 |
| 3 | 42 | 1.6 |
| 4 | 44 | 1.7 |
| 5 | 4 | 0.2 |
| 7 | 71 | 2.7 |
| 9 | 2 | 0.1 |
| 11 | 11 | 0.4 |

**Note.** Total N corresponds to full Sample 2 N (2,613) prior to creating analytic sample.

**Supplementary Table 2. Missingness pattern by domain (restricted to analytic sample, N = 2,525)**

| **Domain** | **Frequency** | **%** |
| --- | --- | --- |
| Executive Function |  |  |
| *2 of 2 tests completed* | 2,353 | 93.2 |
| *1 of 2 tests completed* | 160 | 6.3 |
| *Both tests missing* | 12 | 0.5 |
| Processing Speed |  |  |
| *2 of 2 tests completed* | 2,351 | 93.1 |
| *1 of 2 tests completed* | 164 | 6.5 |
| *Both tests missing* | 10 | 0.4 |
| Episodic Memory |  |  |
| *3 of 3 tests completed* | 2,401 | 95.1 |
| *2 of 3 tests complete* | 58 | 2.3 |
| *1 of 3 tests complete* | 66 | 2.6 |
| *All three tests missing* | 0 | 0.0 |
| Working Memory |  |  |
| *2 of 2 tests completed* | 2,380 | 94.2 |
| *1 of 2 tests completed* | 143 | 5.7 |
| *Both tests missing* | 2 | 0.1 |
| Language/Semantic Fluency |  |  |
| *2 of 2 tests completed* | 2,405 | 95.2 |
| *1 of 2 tests completed* | 55 | 2.2 |
| *Both tests missing* | 65 | 2.6 |

**Section 3: Constructed Measures**

**Supplementary Table 3. Constructed measures used in testing criterion validity of factor scores**

| **Constructed measure** | **Items** | **Coding** |
| --- | --- | --- |
| TestMyBrain (TMB) Summary Score | 1. TMB Verbal Paired Associates (accuracy) | - Each outcome is z-scored (mean = 0, SD = 1) within the analytic sample. - Average of z-scored outcomes serves as the summary score. - Higher scores indicate better performance on TMB. |
|  | 2. TMB Digit Symbol Matching (median reaction time, reverse-coded) |  |
|  | 3. TMB GCPT (dPrime) |  |
|  | 4. TMB Backward Digit Span (span) |  |
| Self-reported memory | 1. I am good at remembering names. | - Each item is scored on a Likert scale 1 to 4 (strongly agree to strongly disagree) - All items summed to create a continuous composite score (range 1 – 20). - Higher values indicate worse self-reported memory. |
|  | 2. I can remember things as well as always. |  |
|  | 3. After I have read a book, I have no difficulty remembering information from it. |  |
|  | 4. The older I get the harder it is to remember clearly (reverse-coded). |  |
|  | 5. Sometimes I have trouble remembering where I have put things (reverse-coded). |  |
| Self-reported hearing | Which statement best describes your hearing without a hearing aid or other assistive devices? | - Item is scored from 1 – 6 (excellent; good; a little trouble; moderate hearing trouble; a lot of trouble; deaf). - Higher values indicate worse self-reported hearing. |
| Self-reported physical function | 1. Walking several blocks | - Each item is scored on a scale of 0 – 2 (no (difficulty), yes, can’t do this). - All items summed to create a continuous composite score (range 0 – 18). - Higher values indicate worse self-reported physical function. |
|  | 2. Sitting for about two hours |  |
|  | 3. Getting up from a chair after sitting for long periods |  |
|  | 4. Climbing several flights of stairs without resting |  |
|  | 5. Stooping, kneeling, or crouching |  |
|  | 6. Reaching or extending your arms above shoulder level. |  |
|  | 7. Pulling or pushing large objects like a living room chair. |  |
|  | 8. Lifting or carrying weights over 10 pounds, like a heavy bag of groceries. |  |
|  | 9. Picking up a dime from a table |  |
| Self-reported health | In general, how is your health? | - Item is scored from 1 – 5 (excellent; very good; good; fair; poor). - Higher values indicate worse self-reported health. |

**Section 4: Factor Diagrams**

Standardized loadings are presented for all models.

**Supplementary Figure 2. Factor diagrams of single-factor models (five domains)**

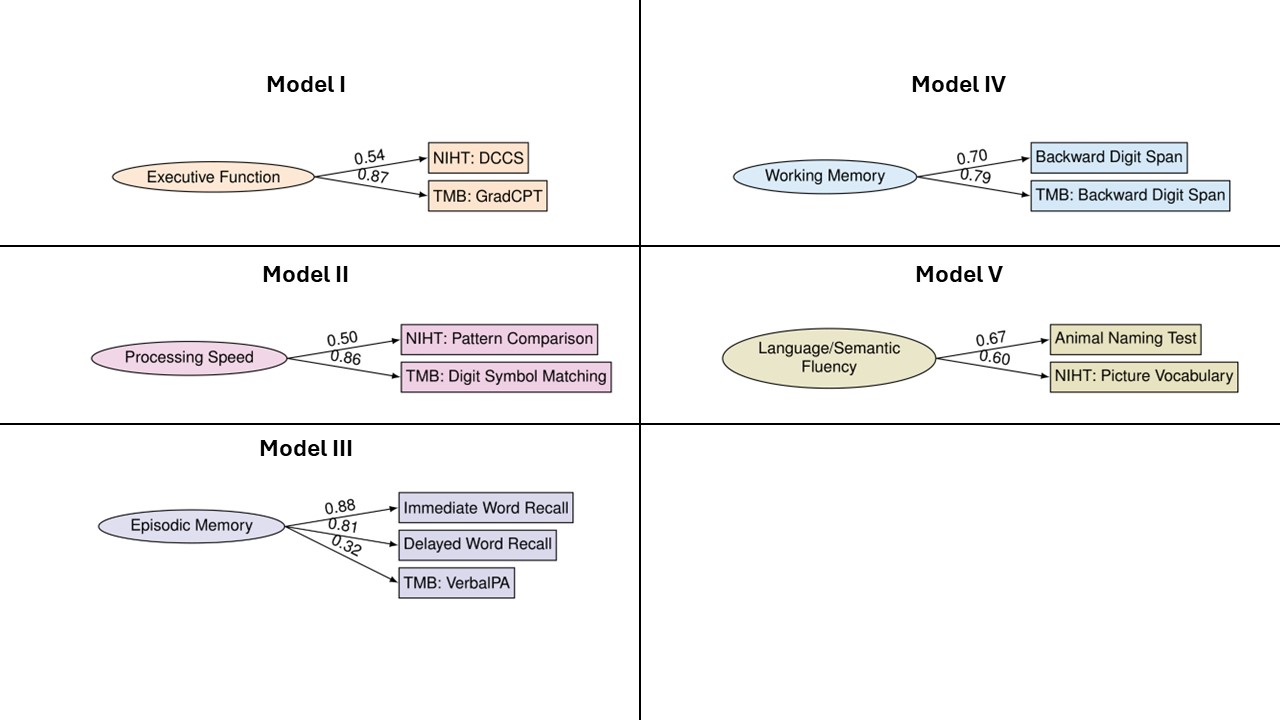

**Supplementary Figure 2 Caption.**

All single-factor models exhibited perfect fit statistics, due to being saturated (degrees of freedom = 0).

**Supplementary Figure 3. Factor diagrams of five-domain correlated factors model**

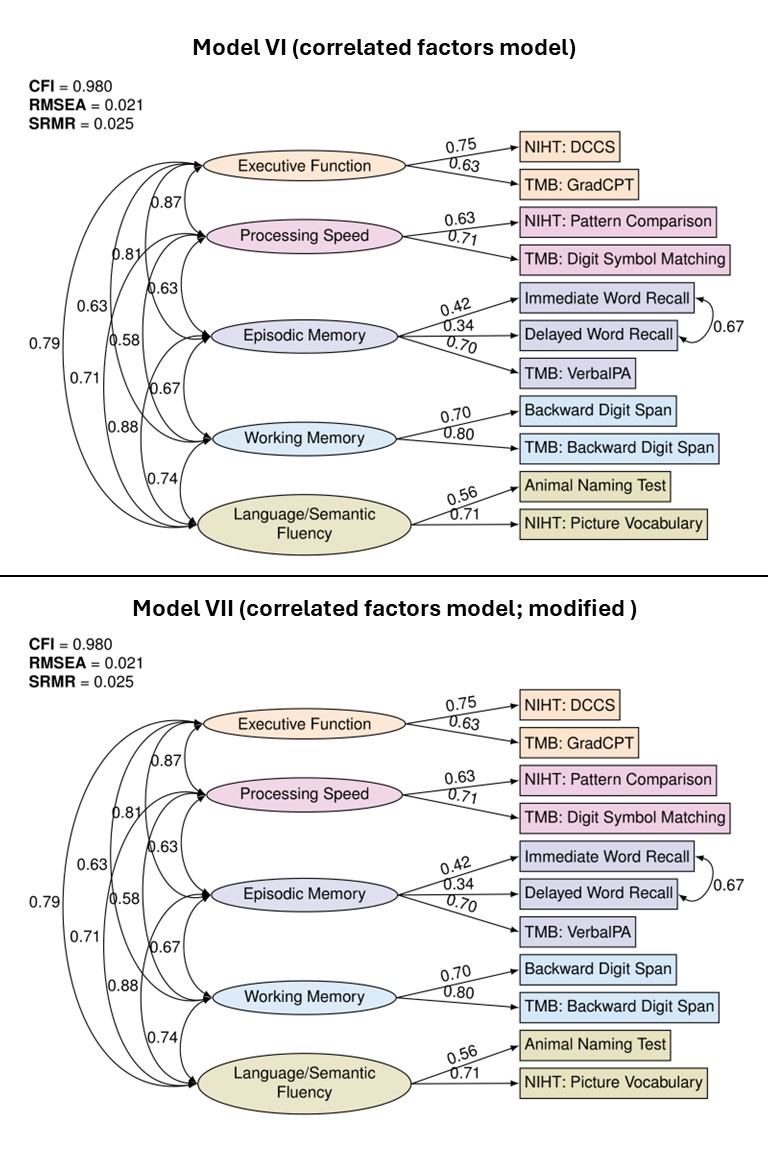

**Supplementary Figure 3 Caption.**

Model VI is the initial correlated factors model conducted, which exhibited poor fit. To improve model fit, residual errors between Immediate and Delayed Word Recall were allowed to correlate, to account for shared variance due to methodological similarities.

**Supplementary Figure 4. Factor diagrams of broad domain of executive function**

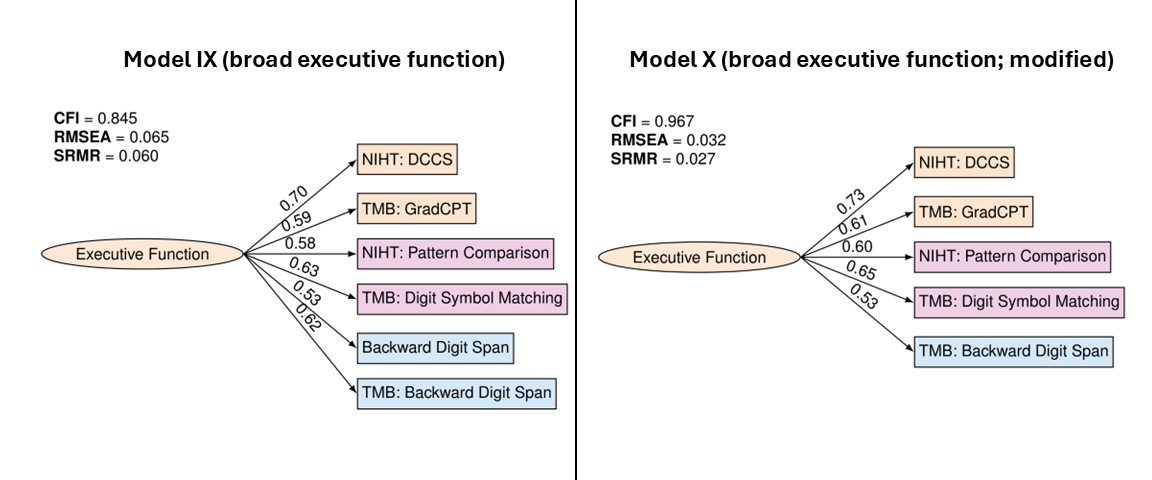

**Supplementary Figure 4 Caption.**

Single-factor models for modified, broad domain of executive function (comprised of executive function, processing speed, and working memory assessments). Model IX displayed poor fit due to redundancy in two Backward Digit Span assessments. Model X shows improved fit, after omission of interviewer-administered Backward Digit Span.

**Supplementary Figure 5. Factor diagram of correlated three-factor model**

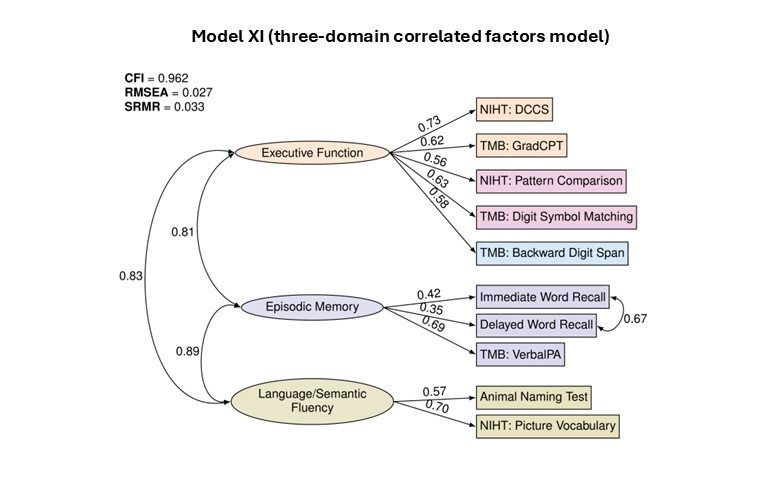

**Section 5: Factor Scores and Criterion Validity**

**Supplementary Figure 6. Distributions of factor scores**

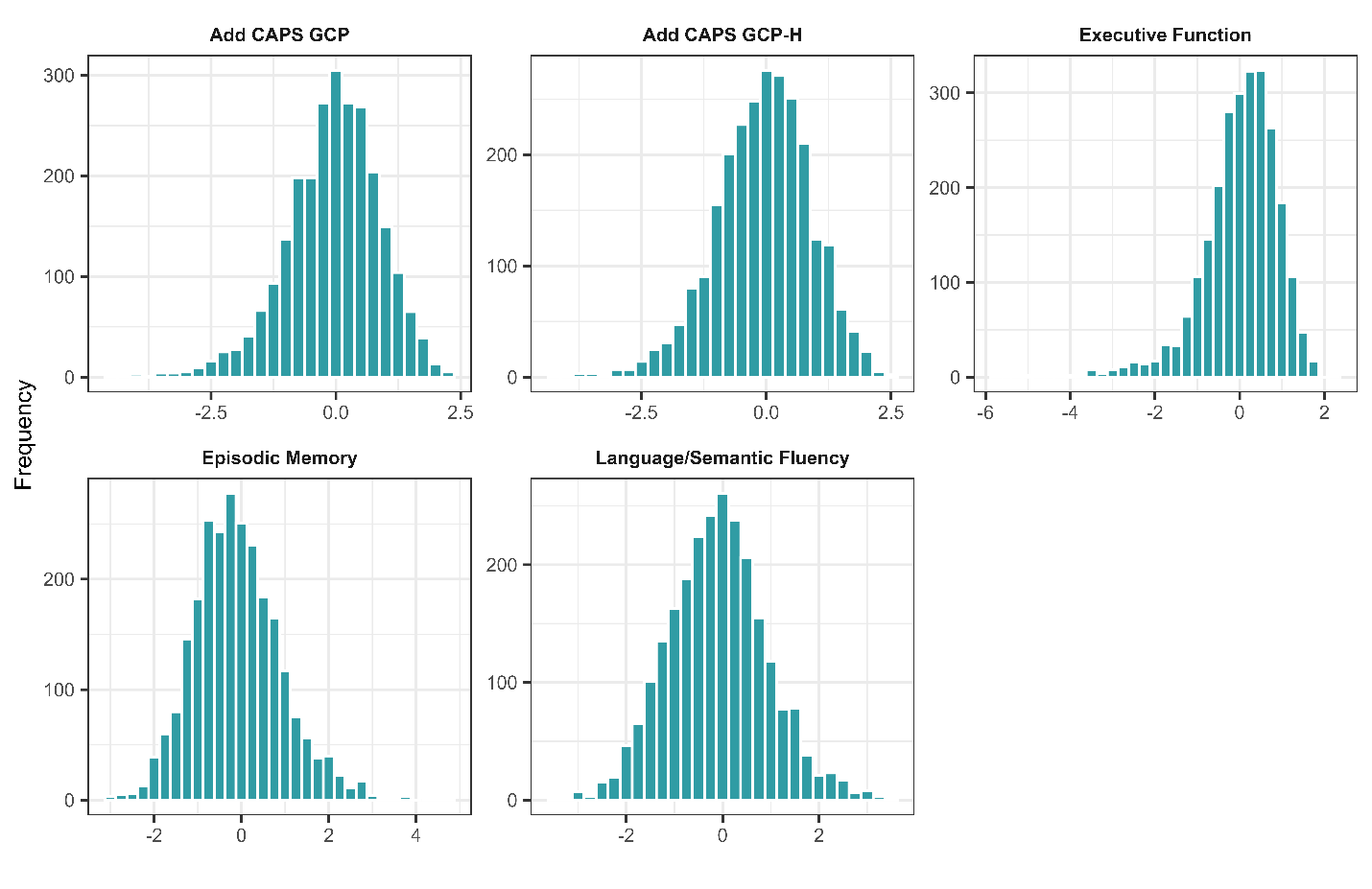

**Supplementary Figure 6 Caption.**

Histograms of factor scores of Add CAPS GCP (Model VIII, N = 2,525), Add CAPS GCP-H (Model XII, N = 2,525), executive function (Model X, N = 2,516), episodic memory (Model III, N = 2,525) and language/semantic fluency (Model V, N = 2,460) are presented. All scores were standardized to a mean of 0, and standard deviation of 1.

**Supplementary Figure 7. Survey-weighted Pearson’s correlations among cognition factor scores and sociodemographic and health variables**

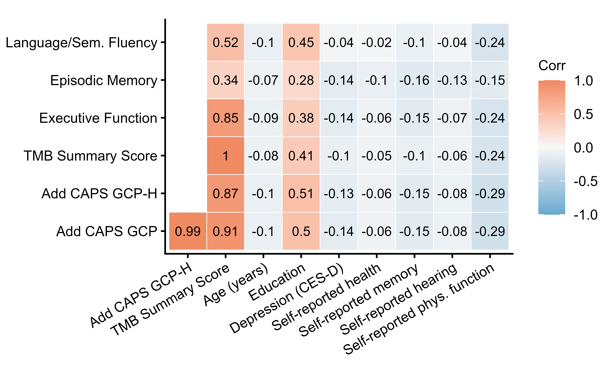

**Supplementary Figure 7 Caption.**

TMB Summary Score is the average of four z-scored TMB assessment outcomes. Self-reported health, memory, hearing, and physical function = higher values indicate worse self-reported health, memory, hearing, and physical function. Education refers to the highest level of education completed (values range from 1: “8^th^ grade or less” to 16: “completed a post baccalaureate professional degree (such as law, medicine, nursing)”).
